# Is scaling-up COVID-19 testing cost-saving?

**DOI:** 10.1101/2020.03.22.20041137

**Authors:** Bernardo Sousa-Pinto, João Almeida Fonseca, Altamiro Costa-Pereira, Francisco Nuno Rocha-Gonçalves

## Abstract

The World Health Organization currently recommends that governments scale up testing for COVID-19 infection. We performed health economic analyses projecting whether the additional costs from screening would be offset by the avoided costs with hospitalizations. We analysed Portuguese COVID-19 data up until the 22nd March 2020, and estimated the additional number of cases that would be detected if different testing rates and frequencies of positive results would have been observed. We projected that, in most scenarios, the costs with scaling up COVID-19 tests would be lower than savings with hospitalization costs, rendering large scale testing cost-saving.

---

The World Health Organization currently recommends that governments scale up testing for COVID-19 infection[1]. Large-scale testing is frequently pointed out as one of the reasons behind South Korea success in dealing with COVID-19 infection[2]. Costs may be regarded as an important barrier for scaling up testing. Nevertheless, earlier detection of cases subsequent to large-scale testing may prevent new infections, prompting savings in hospitalizations. Therefore, we performed health economic analyses aiming to assess whether, in Portugal, scaling up testing could be cost-saving.

Portugal is a European country with 10.3 million inhabitants[3]. Until 22^nd^ March 2020, 10,627 individuals had been tested for COVID-19, with 1600 (15.1%) positive results[4]. This corresponds to 1034.1 tests and 155.8 cases per million inhabitants. We calculated the additional number of cases that would be detected if different testing rates and frequencies of positive results would have been observed. In our base-case models, we considered that, by earlier detection of such additional cases (*a*), we would be preventing *a*(1+0.29)^10^ new infections over the following 10 days (considering an infection growth rate of 29.0%, corresponding to the Portuguese average of the five previous days, and the exponential growth formula). We considered the National Health Service (NHS) perspective for costs. In our base-case models, we assumed testing costs of 150 Euro *per* patient (corresponding to processing three samples with molecular tests at 50 Euro each[5]). Savings in additional hospitalizations were estimated according to the inputs and assumptions displayed in Table 1.

**Table 1.** Model inputs and respective sources

| Variable | Mean | Range | Median | Proportion | Distribution | Source |
| --- | --- | --- | --- | --- | --- | --- |
| Number of tested patients per million inhabitants | - | 900-3000 | - | - | <sup>a</sup> | - |
| Frequency of positive test results | - | 0-0.25 | - | - | <sup>a</sup> | - |
| Daily COVID-19 infection growth rate | 0.29 | 0.22-0.43 | - | - | Triangular | <sup>b</sup> |
| Costs of COVID-19 testing *per* patient (Euro) | - | 75-150 | - | - | <sup>a</sup> | <sup>c</sup> |
| Frequency of hospitalized COVID-19 patients | - | - | - | 0.106 | Beta | <sup>d</sup> |
| Frequency of hospitalized COVID-19 patients receiving care in ICU | - | - | - | 0.243 | Beta | <sup>d</sup> |
| Length of stay of COVID-19 hospitalizations (days) | 14 | - | 12 | - | Lognormal | <sup>e</sup> |
| Daily costs of a COVID-19 admission (Euro) | 338.9 | - | 209.5 | - | Lognormal | <sup>f</sup> |
| Daily costs of an ICU admission (Euro) | 1174.3 | - | 823.0 | - | Lognormal | <sup>f</sup> |

ICU=Intensive care unit; <sup>a</sup> Fixed values; <sup>b</sup> Input values chosen based on the Portuguese average, minimum and maximum daily infection growth rates of the five previous days as according to official sources (*Direção Geral da Saúde* - https://www.dgs.pt/); <sup>c</sup> The base case value assumed a cost of 150 Euro for testing one patient (corresponding to three samples costing 50 Euro each); <sup>d</sup> Communication of the Health General Direction (*Direção Geral da Saúde*) as noticed in https://www.publico.pt/2020/03/22/sociedade/noticia/portugal-1600-casos-coronavirus-1908904; <sup>e</sup> Open data from Xu B et al. Lancet Infectious Diseases. 2020 (doi.org/10.1016/S1473-3099(20)30119-5); <sup>f</sup> Portuguese database of hospital admissions – we considered the daily costs of hospitalizations by pneumonia as indicative of the daily costs of COVID-19 admissions (DRG=139 as defined by the Portuguese Ministry of Health in http://www2.acss.min-saude.pt/Default.aspx?TabId=922&language=pt-PT). Costs of pneumonia hospitalizations requiring ventilator use >96 hours were used to estimate daily costs of COVID-19 admissions in intensive care units (DRG=130 as defined by the Portuguese Ministry of Health in http://www2.acss.min-saude.pt/Default.aspx?TabId=922&language=pt-PT).

We projected whether there would be net economic savings or losses if different testing rates and frequencies of positive results would have been observed – several combinations of predefined values were analysed. For each combination, we performed probabilistic sensitivity analyses via Monte Carlo simulations, estimating the percentage of simulations identifying a change in testing as cost-saving. In addition, we performed sensitivity analyses varying the costs of testing, to account for possible future decreases in testing costs. We compared the results of our models with the scenarios in other four countries by the date they reported a similar number of cases per million inhabitants.

Model results are presented in Table 2 and Figures 1-2. Our results project that scaling up COVID-19 tests would be net cost-saving (i.e., costs lower than savings with hospitalization costs) in most scenarios, particularly when the frequency of positive tests does not reach excessively low values.

**Table 2.**
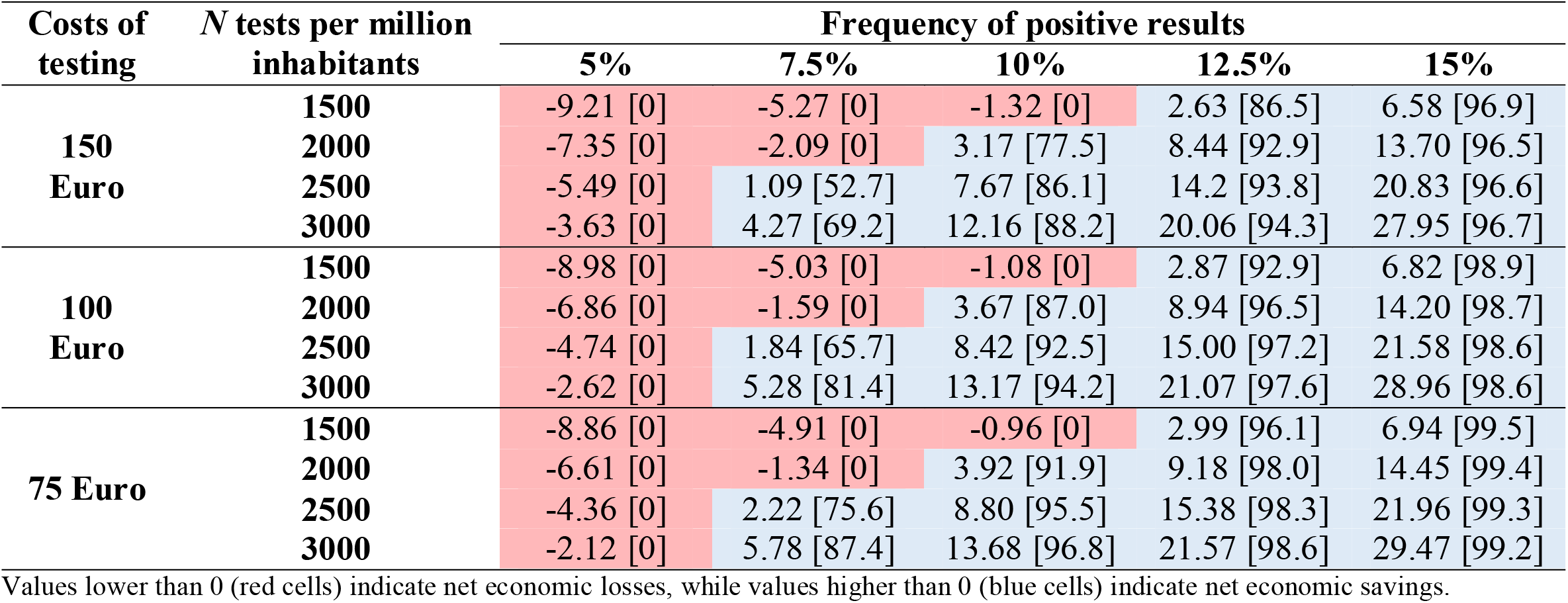
Projected economic savings or losses that would be observed under different combinations of number of COVID-19 tests per million inhabitants and frequency of positive test results. Results are presented in million Euro, along with the percentage of probabilistic sensitivity analyses identifying changes in testing strategies as cost-saving (in square brackets)

| Costs of testing | N tests per million inhabitants | Frequency of positive results |  |  |  |  |
| --- | --- | --- | --- | --- | --- | --- |
|  |  | 5% | 7.5% | 10% | 12.5% | 15% |
| **150 Euro** | **1500** | -9.21 [0] | -5.27 [0] | -1.32 [0] | 2.63 [86.5] | 6.58 [96.9] |
|  | **2000** | -7.35 [0] | -2.09 [0] | 3.17 [77.5] | 8.44 [92.9] | 13.70 [96.5] |
|  | **2500** | -5.49 [0] | 1.09 [52.7] | 7.67 [86.1] | 14.2 [93.8] | 20.83 [96.6] |
|  | **3000** | -3.63 [0] | 4.27 [69.2] | 12.16 [88.2] | 20.06 [94.3] | 27.95 [96.7] |
| **100 Euro** | **1500** | -8.98 [0] | -5.03 [0] | -1.08 [0] | 2.87 [92.9] | 6.82 [98.9] |
|  | **2000** | -6.86 [0] | -1.59 [0] | 3.67 [87.0] | 8.94 [96.5] | 14.20 [98.7] |
|  | **2500** | -4.74 [0] | 1.84 [65.7] | 8.42 [92.5] | 15.00 [97.2] | 21.58 [98.6] |
|  | **3000** | -2.62 [0] | 5.28 [81.4] | 13.17 [94.2] | 21.07 [97.6] | 28.96 [98.6] |
| **75 Euro** | **1500** | -8.86 [0] | -4.91 [0] | -0.96 [0] | 2.99 [96.1] | 6.94 [99.5] |
|  | **2000** | -6.61 [0] | -1.34 [0] | 3.92 [91.9] | 9.18 [98.0] | 14.45 [99.4] |
|  | **2500** | -4.36 [0] | 2.22 [75.6] | 8.80 [95.5] | 15.38 [98.3] | 21.96 [99.3] |
|  | **3000** | -2.12 [0] | 5.78 [87.4] | 13.68 [96.8] | 21.57 [98.6] | 29.47 [99.2] |

Values lower than 0 (red cells) indicate net economic losses, while values higher than 0 (blue cells) indicate net economic savings.

**Figure 1.**
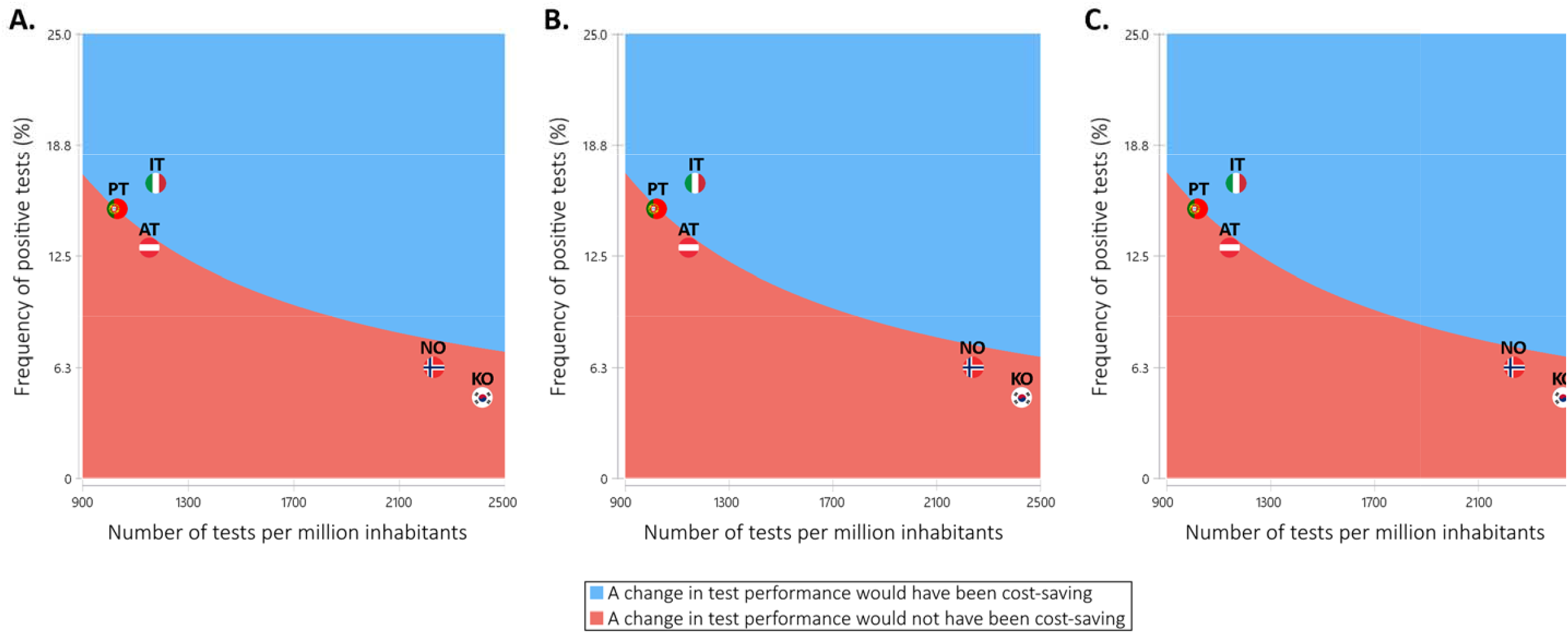
Results of the models assessing whether a change in COVID-19 test performance would result be cost-saving (i.e., whether the costs of testing would be lower than savings with hospitalization costs), and assuming that testing each patient costs 150 Euro (A), 100 Euro (B), or 75 Euro (C). AT=Austria (data until 17^th^ March 2020 – 149.6 cases per million inhabitants); IT=Italy (data until 10^th^ March 2020 – 168.3 cases per million inhabitants); KO=South Korea (data until 5^th^ March 2020 – 111.7 cases per million inhabitants); NO=Norway (data until 13^th^ March 2020 – 139.7 cases per million inhabitants); PT=Portugal (data until 22^nd^ March 2020 – 155.8 cases per million inhabitants)

**Figure 2.**
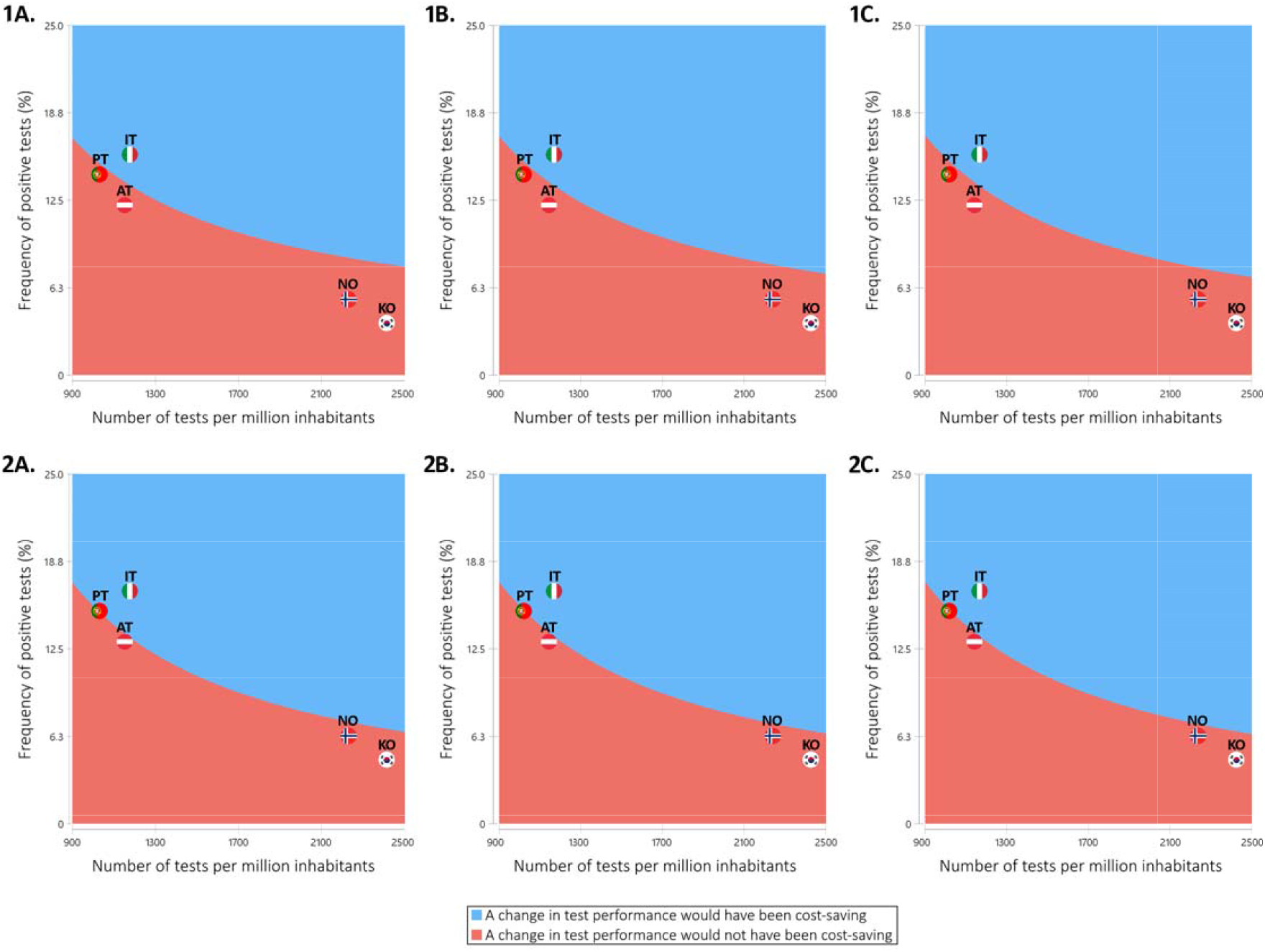
Results of sensitivity analysis models assessing whether a change in COVID-19 test performance would result in lower costs, assuming (1) a daily infection growth rate of 22%, and (2) a daily infection growth rate of 43%.Costs of testing each patient are being assumed as corresponding to 150 Euro (A), 100 Euro (B), and 75 Euro (C). AT=Austria (data until 17^th^ March 2020 – 149.6 cases per million inhabitants); IT=Italy (data until 10^th^ March 2020 – 168.3 cases per million inhabitants); KO=South Korea (data until 5^th^ March 2020 – 111.7 cases per million inhabitants); NO=Norway (data until 13^th^ March 2020 – 139.7 cases per million inhabitants); PT=Portugal (data until 22^nd^ March 2020 – 155.8 cases per million inhabitants)

To keep an adequate frequency of positive tests, there is need for data (i) to better define testing criteria, and (ii) to understand the preanalytical and analytical vulnerabilities of COVID-19 testing[6].

This study has important inherent limitations, as the built models are, necessarily, oversimplifications of reality. Also, data is still scarce – for instance, the number of cases that are currently undiagnosed cannot be yet known, and there are limited available accurate numbers of performed COVID-19 tests in many countries. Nevertheless, we have chosen a conservative approach, possibly underestimating hospitalizations costs – the daily costs of COVID-19 admissions are probably higher than those established for pneumonia. In addition, other relevant costs in the NHS perspective are not being considered, as they would be difficult to measure – because hospitals are being directed to treat COVID-19 infections, there can be postponed treatments, and drugs/technologies where COVID-19 patients may be given priority over other patients. Thus, real savings subsequent to large scale testing may be higher than those we presented, as we did not consider opportunity costs and indirect costs (e.g., productivity losses subsequent to admissions or quarantines).

Although these are short-term models, the ever-changing nature of COVID-19 pandemic and the dynamic nature of the variables of this study – for example the development of cheaper and faster diagnostic kits – may prompt large-scale testing to become even more appealing.

In fact, the cost COVID-19 tests is expected to vary, as companies are quickly bringing to the market new technologies that promise automated processing and faster response rates. Of note, we are only considering molecular tests, and not serological ones, which, being cheaper, are not currently considered adequate for diagnostic purposes. Value of diagnostics usually encompasses turnover time, and both regulators and industry promoters are addressing this topic as well[7]. Therefore, test prices will likely be pressured downwards, mirroring more offer coming into the market and the regulatory pathways that support this rapid market entry[8].

In summary, this study, highlights the potential net cost-saving effects of scaling up COVID-19 testing, supporting the importance of building up test capacity in the health system at the earliest possible time so that more hospitalizations can be prevented, resulting in lower pressure on the healthcare system and better outcomes for patients.

## Data Availability

Data used to build the economic models can be provided on request.

## Notes

### Competing Interest Statement

The authors have declared no competing interest.

### Funding Statement

No external funding was received.

## References

1. Offord C. Governments must ramp up COVID-10 Testing, says WHO. The Scientist. 18^th^ March 2020. (https://www.the-scientist.com/news-opinion/governments-must-ramp-up-covid-19-testing-says-who-67287) [last accessed – 21st March 2020].

2. Cohen J, Kupferschmidt K. Countries test tactics in ‘war’ against COVID-19. Science. 2020:367(6484):1287–1288.

3. Portuguese National Institute of Statistics [Instituto Nacional de Estatística] (https://www.ine.pt/) [last accessed – 22nd March 2020].

4. Portuguese Health Directorate. COVID-19 – Portuguese epidemiological report updated at 22nd March 2020 [Direcção-Geral da Saúde. COVID-19 - Relatório de Situação. Situação Epidemiológica em Portugal. Actualizado a 22 de Março de 2020]. (https://www.dgs.pt/em-destaque/relatorio-de-situacao-n-020-22032020-pdf.aspx) [last accessed – 22nd March 2020].

5. Centers for Medicare & Medicaid Services. Medicare Administrative Contractor COVID-19 Test Pricing March 12, 2020 (https://www.cms.gov/files/document/mac-covid-19-test-pricing.pdf) [last accessed – 22nd March 2020].

6. Lippi G, Simundic AM, Plebani M. Clin Chem Lab Med. 2020. Doi:10.1515/cclm-2020-0285.

7. Food and Drug Administration. Emergency Use Authorizations: Coronavirus Disease 2019 (COVID-19) Emergency Use Authorizations for Medical Devices (https://www.fda.gov/medical-devices/emergency-situations-medical-devices/emergency-use-authorizations#coronavirus2019) [last accessed – 22nd March 2020].

8. Food and Drug Administration. Policy for Diagnostic Tests for Coronavirus Disease-2019 during the Public Health Emergency (https://www.fda.gov/regulatory-information/search-fda-guidance-documents/policy-diagnostic-tests-coronavirus-disease-2019-during-public-health-emergency) [last accessed – 22nd March 2020].

